# Peripartum cardiomyopathy is independently associated with pre-existing cardiovascular disease and smoking, with Loss of Racial Disparities After Multivariate Adjustment

**DOI:** 10.64898/2025.12.12.25342183

**Authors:** Logan T. Kenny, Sumedha Tadimeti, Mehrtash Hashemzadeh, Mohammad Reza Movahed

## Abstract

**Background:** Peripartum cardiomyopathy (PPCM) is a life-threatening condition. Despite rising incidence, outcomes have not improved. This study evaluated national inpatient trends, disparities, and independent risk factors for PPCM.

**Methods:** Using the National Inpatient Sample (2016–2020), we analyzed adults (≥18 years) hospitalized between 10 and 42 weeks of gestation. Among 18,844,715 patients, 4,475 had PPCM. We performed uni- and multivariate logistic regression analysis of risk factors for PCCM, adjusting for age, race, comorbidities, hospital characteristics, and socioeconomic factors.

**Results:** PPCM was associated with higher maternal mortality (1.01% vs 0.01%; OR 164.41, 95% CI 84.17–321.12). Univariate analysis showed a higher association with preeclampsia, gestational diabetes, gestational hypertension, preexisting hypertension, coronary artery disease (CAD), chronic kidney disease, systemic lupus erythematosus, type 2 diabetes, obesity and smoking. PPCM was more common in Black and Native American patients, those younger than 30, and individuals from lower-income households. However, after adjustment, significant associations only persisted for preeclampsia, gestational hypertension, preexisting hypertension, CAD, and smoking. CAD showed the strongest association in both univariate (OR 125.86, 95% CI 89.17–177.66) and multivariate models (OR 3.29, 95% CI 2.19–4.95). Racial disparities were no longer significant after adjustment.

**Conclusion:** PPCM is associated with higher mortality. It is independently associated with preexisting cardiovascular risk factors. Apparent racial disparities primarily reflect comorbidities and social determinants and are not independent risk.

## Introduction

Peripartum cardiomyopathy (PPCM) is a life-threatening cardiac disease that classically arises during late pregnancy or early postpartum and is characterized by heart failure due to left ventricular systolic dysfunction ^1^. It was reported as early as 1849 ^2^ but was not identified as a distinct diagnosis until the 1930s ^3^.

The prevailing view is that the mechanism is multifactorial. Potential contributors include autoimmune phenomena, inadequate nutrition, the hemodynamic burden of pregnancy, and hormonally driven vascular insult in patients with underlying genetic susceptibility ^4^. The pathophysiological mechanism underlying this disease is postulated to be from multiple factors such as mutations in sarcomeric and DNA-repair genes, hypertension, and heightened β-adrenergic activity, which can amplify oxidative stress, leading to cleavage of prolactin into a 16-kDa N-terminal fragment. This fragment impairs the vasculature by causing endothelial dysfunction and endothelial cell death. The resulting reduction in oxygen and nutrient delivery, along with diminished cardiomyocyte metabolic capacity, ultimately contributes to the development of peripartum cardiomyopathy ^5^. Interestingly, A 2021 study of patients with peripartum cardiomyopathy demonstrated substantial genetic overlap with dilated cardiomyopathy. It also identified pathogenic truncating variants in the TTN gene in approximately 10 percent of women affected by PPCM, representing about a 20-fold higher prevalence compared with control cohorts ^6^.

There appears to be a regional difference in PPCM diagnoses as incidence is highest in countries with tropical climates and distinct wet and dry seasons, ranging from 1 per 102 live births in Nigeria to 1 per 299 in Haiti. Other nations, including Pakistan, the Philippines, China, Sudan, and South Africa, report approximately 1 case of PPCM per 1000 live births ^5^. Anti-inflammatory therapy with pentoxifylline, a xanthine derivative that inhibits TNF and IL-6 release, has been shown to improve cardiac function and clinical outcomes in an African cohort with PPCM ^7^. The source of the heightened inflammatory state in these patients remains unclear and may relate to autoimmune mechanisms that evolved to protect pregnant women in tropical regions. This all goes to highlight the importance of assessing risk in terms of regional differences.

A previous national study in the United States identified an increase in PPCM incidence from 8.5 to 11.8 (average of 10.3) diagnoses per 10,000 live births from 2004 to 2011 ^8^. Despite the increase in incidence, the prognosis of PPCM is not improving ^9^. The lack of improvement in care may be due to the difficulty of distinguishing between PPCM symptoms and the normal symptoms that arise during pregnancy ^10^. Of additional note, although many women with peripartum cardiomyopathy present with moderate to severe symptoms (NYHA Class III or IV), a study noted that there is a poor correlation between PPCM symptoms and LVEF. In fact, a quarter of those with mild (NYHA I/II) symptoms had a LVEF ≤25% ^11^. Collectively, these facts make identifying risk factors and epidemiological trends in PPCM all the more important. Several risk factors have been identified in association with PPCM diagnoses. A recent systematic review and meta-analysis found risk factors to include obesity, multiparity, gestational hypertension, diabetes, and preeclampsia ^12^. It has also been reported that this disease is four times more likely to develop in Black women compared to White women in the United States ^13^. While previous studies have identified several risk factors, data specific to U.S. inpatient settings with thorough adjustment remain limited. The goal of our study was to evaluate independent risk factors for PPCM.

## Methods

We evaluated adult females (≥18 years) hospitalized between 10 and 42 weeks of gestation from 2016 to 2020. We assessed a range of demographic variables, outcomes, complications, and risk factors identified via ICD-10 codes (Table S1). Continuous variables were summarized using means with standard deviations (SD) or medians with interquartile ranges (IQR), as appropriate. Differences in continuous variables between patients with and without PPCM were assessed using independent samples t-tests for normally distributed variables and Mann–Whitney U tests for non-normally distributed variables. Categorical variables were compared using Chi-square tests.

### Univariate Models

Univariate logistic regression models were used to estimate unadjusted odds ratios (ORs) with 95% confidence intervals (CIs) for the association between PPCM (dependent variable) and each clinical outcome, including mortality, pregnancy-associated complications, and chronic comorbidities.

### Multivariate (Adjusted) Models

Variables with p<0.001 in univariate models were included in multivariate logistic regression analyses to adjust for potential confounders, including age, race, comorbid conditions, hospital characteristics, and socioeconomic factors, when assessing the association with PPCM (dependent variable). All p-values were 2-sided, with p<0.05 considered statistically significant. Analyses were performed using STATA 17 (Stata Corporation, College Station, TX).

## Results

### Univariate Models

#### General

Among 18,844,715 pregnant patients hospitalized from 2016–2020, 4,475 (0.02%) were diagnosed with PPCM. Women with PPCM were older on average (30.40 ± 6.12 years) than those without PPCM (29.09 ± 5.69 years, p < 0.001). Maternal mortality was markedly higher in the PPCM group (1.01% vs 0.01%; OR 164.41, 95% CI 84.17–321.12, p < 0.001) (Table 1).

**Table 1.** Univariate Analysis – *Sourced from O10 (“preexisting hypertension complicating pregnancy”), which is more complete in pregnant inpatients than general chronic hypertension codes.

|  | **PPCM (%)** | **No PPCM (%)** | **OR** | **95% CI** | **p-value** |
| --- | --- | --- | --- | --- | --- |
| **Maternal mortality** | 1.01 | 0.01 | 164.41 | 84.17–321.12 | <0.001 |
| **Pregnancy-Associated Conditions** |  |  |  |  |  |
| Eclampsia | 1.68 | 0.07 | 23.12 | 13.85–38.61 | <0.001 |
| Preeclampsia | 37.77 | 6.16 | 9.25 | 8.05–10.62 | <0.001 |
| Spontaneous abortion | 0.78 | 0.14 | 5.81 | 2.76–12.24 | <0.001 |
| Gestational edema/proteinuria (no hypertension) | 1.56 | 0.39 | 4.01 | 2.37–6.80 | <0.001 |
| Hemorrhage in early pregnancy | 0.22 | 0.07 | 3.04 | 0.76–12.20 | 0.12 |
| Genitourinary tract infection in pregnancy | 3.69 | 1.30 | 2.91 | 2.07–4.09 | <0.001 |
| Gestational diabetes | 15.87 | 9.45 | 1.81 | 1.51–2.16 | <0.001 |
| Gestational hypertension (no proteinuria) | 8.94 | 5.94 | 1.55 | 1.23–1.96 | <0.001 |
| Excessive vomiting in pregnancy | 0.89 | 0.78 | 1.14 | 0.57–2.29 | 0.70 |
| Malnutrition in pregnancy, childbirth and the puerperium | 1.23 | 0.04 | 28.51 | 15.72–51.70 | <0.001 |
| **Chronic Medical Conditions** |  |  |  |  |  |
| Hypertension (preexisting)* | 17.32 | 3.08 | 6.59 | 5.57–7.81 | <0.001 |
| Coronary artery disease (CAD) | 3.91 | 0.03 | 125.86 | 89.17–177.66 | <0.001 |
| Chronic kidney disease (CKD) | 2.57 | 0.18 | 14.76 | 9.60–22.70 | <0.001 |
| Chronic obstructive pulmonary disease (COPD) | 1.12 | 0.05 | 21.48 | 11.47–40.22 | <0.001 |
| Hyperlipidemia | 2.79 | 0.36 | 7.95 | 5.35–11.82 | <0.001 |
| Lupus (SLE) | 1.12 | 0.16 | 7.00 | 3.76–13.04 | <0.001 |
| Type 2 diabetes | 4.47 | 0.88 | 5.26 | 3.86–7.18 | <0.001 |
| Rheumatoid arthritis | 0.45 | 0.13 | 3.35 | 1.26–8.94 | 0.02 |
| Obesity | 11.06 | 6.18 | 1.89 | 1.54–2.33 | <0.001 |
| Smoking | 12.96 | 6.73 | 2.06 | 1.70–2.51 | <0.001 |
| Alcohol use | 0.22 | 0.16 | 1.36 | 0.34–5.47 | 0.66 |

### Pregnancy-Associated Conditions

Compared to women without PPCM, those with PPCM had higher odds of eclampsia (OR 23.12, 95% CI 13.85–38.61), preeclampsia (OR 9.25, 95% CI 8.05–10.62), spontaneous abortion (OR 5.81, 95% CI 2.76–12.24), gestational edema/proteinuria without hypertension (OR 4.01, 95% CI 2.37–6.80), genitourinary tract infection (OR 2.91, 95% CI 2.07–4.09), gestational diabetes (OR 1.81, 95% CI 1.51–2.16), gestational hypertension without proteinuria (OR 1.55, 95% CI 1.23–1.96), and malnutrition in pregnancy (OR 28.51, 95% CI 15.72–51.70) (all p < 0.001). There were no significant differences in hemorrhage in early pregnancy (p = 0.12) or excessive vomiting in pregnancy (p = 0.70) (Table 1).

### Chronic Medical Conditions

Women with PPCM had higher odds of preexisting hypertension (OR 6.59, 95% CI 5.57–7.81), coronary artery disease (OR 125.86, 95% CI 89.17–177.66), chronic kidney disease (OR 14.76, 95% CI 9.60–22.70), chronic obstructive pulmonary disease (OR 21.48, 95% CI 11.47–40.22), hyperlipidemia (OR 7.95, 95% CI 5.35–11.82), systemic lupus erythematosus (OR 7.00, 95% CI 3.76–13.04), type 2 diabetes (OR 5.26, 95% CI 3.86–7.18), rheumatoid arthritis (OR 3.35, 95% CI 1.26–8.94), obesity (OR 1.89, 95% CI 1.54–2.33), and smoking (OR 2.06, 95% CI 1.70–2.51) (all p < 0.05). There was no significant difference in alcohol use (p = 0.66) (Table 1).

### Demographics

In univariate analysis, women with PPCM were disproportionately more likely to be Black or Native American and to come from low-income households (up to $45,999) compared to the overall obstetric population. Despite accounting for only 15.55% of the sampled patients, Black women made up 30.10% of the patients with PPCM (p<0.001). Similarly, Native American patients accounted for 0.75% of the total sample but made up 1.73% of the patients with PPCM (p<0.001). Low-income households accounted for 28.11% of sampled patients, but 40.34% of patients with PPCM (P<0.001). Furthermore, for age, our sample included 41.37% patients <30; however, nearly half (49.72%) of women with PPCM were <30 years old (p<0.001). Those >=30 accounted for 58.63% of the total sample, but only 50.28% of those with PPCM (p<0.001) (Table 2). The hospital region was not significantly different.

**Table 2.**
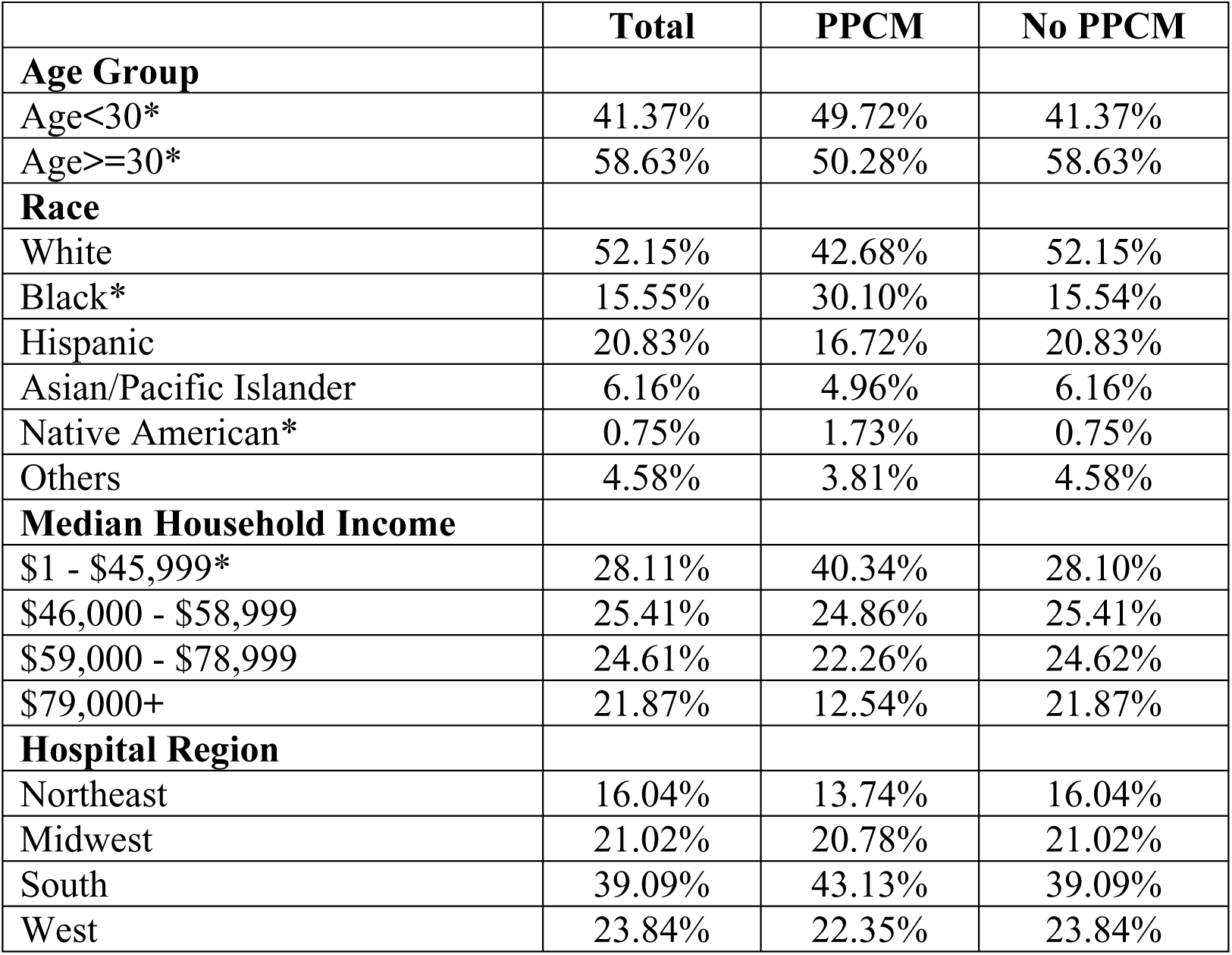
Univariate Demographics – p-value of <0.001 signified by *.

|  | **Total** | **PPCM** | **No PPCM** |
| --- | --- | --- | --- |
| **Age Group** |  |  |  |
| Age<30* | 41.37% | 49.72% | 41.37% |
| Age>=30* | 58.63% | 50.28% | 58.63% |
| **Race** |  |  |  |
| White | 52.15% | 42.68% | 52.15% |
| Black* | 15.55% | 30.10% | 15.54% |
| Hispanic | 20.83% | 16.72% | 20.83% |
| Asian/Pacific Islander | 6.16% | 4.96% | 6.16% |
| Native American* | 0.75% | 1.73% | 0.75% |
| Others | 4.58% | 3.81% | 4.58% |
| **Median Household Income** |  |  |  |
| \$1 - \$45,999* | 28.11% | 40.34% | 28.10% |
| \$46,000 - \$58,999 | 25.41% | 24.86% | 25.41% |
| \$59,000 - \$78,999 | 24.61% | 22.26% | 24.62% |
| \$79,000+ | 21.87% | 12.54% | 21.87% |
| **Hospital Region** |  |  |  |
| Northeast | 16.04% | 13.74% | 16.04% |
| Midwest | 21.02% | 20.78% | 21.02% |
| South | 39.09% | 43.13% | 39.09% |
| West | 23.84% | 22.35% | 23.84% |

### Summary

In summary, women with PPCM were more likely to have eclampsia, preeclampsia, spontaneous abortion, gestational edema or proteinuria without hypertension, genitourinary tract infection, gestational diabetes, gestational hypertension without proteinuria, and malnutrition in pregnancy compared to women without PPCM. Women with PPCM were also more likely to have preexisting hypertension, coronary artery disease, chronic kidney disease, chronic obstructive pulmonary disease, hyperlipidemia, systemic lupus erythematosus, type 2 diabetes, rheumatoid arthritis, obesity, and smoke compared to women without PPCM (Figure 1). Women with PPCM were disproportionately more likely to be Black, Native American, come from low-income households (up to $45,999), or <30 years old compared to the overall obstetric population.

**Figure 1.**
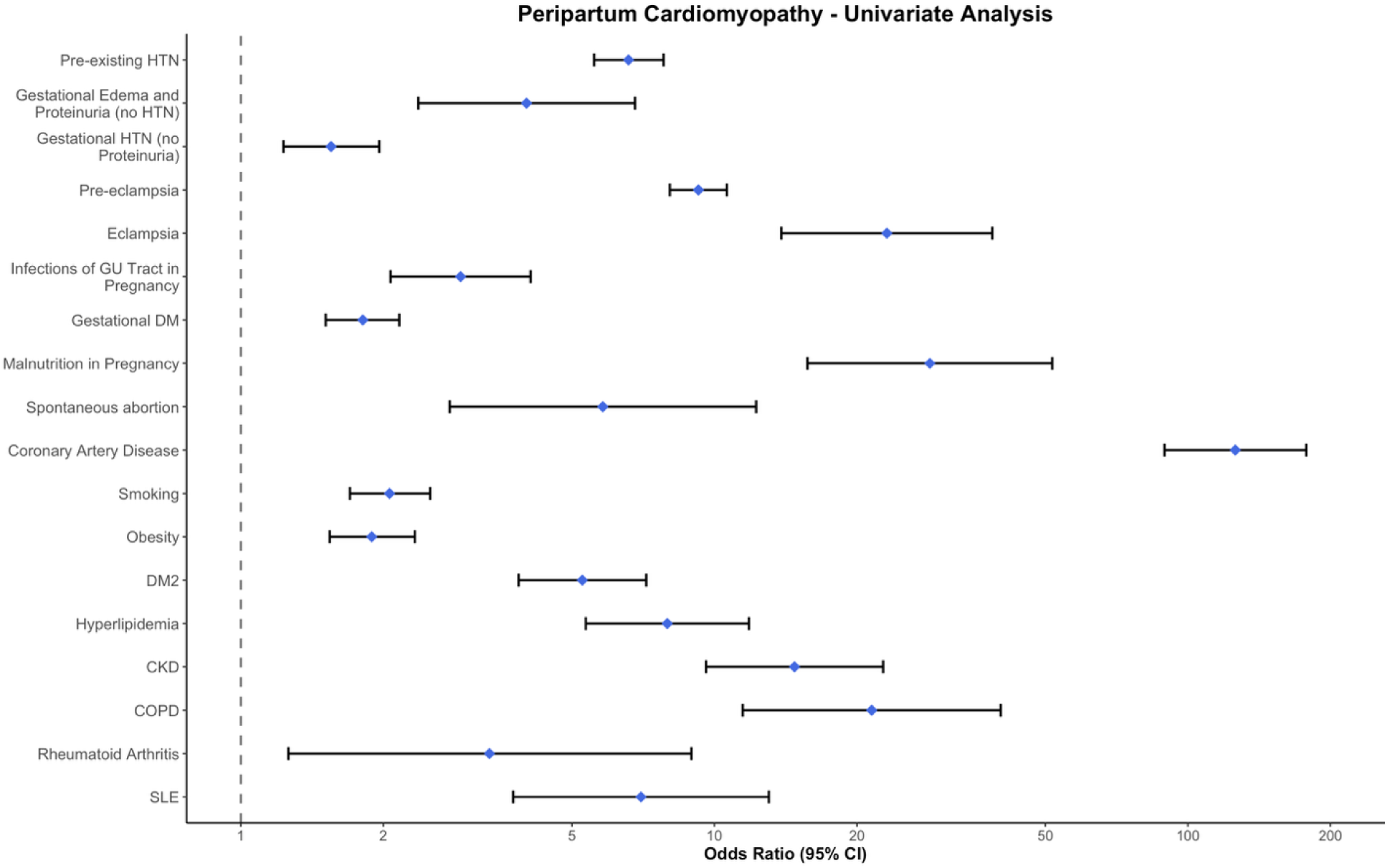
Univariate Significant Results.

### Multivariate (Adjusted) Models

#### Pregnancy-Associated Conditions

After adjustment for age, race, comorbidities, hospital characteristics, and socioeconomic factors, women with PPCM had higher odds of preeclampsia (OR 2.38, 95% CI 2.04–2.78, p < 0.001) and gestational hypertension without proteinuria (OR 1.66, 95% CI 1.31–2.11, p < 0.001). Associations observed in the univariate analysis for eclampsia, spontaneous abortion, gestational edema/proteinuria without hypertension, and malnutrition were no longer statistically significant after adjustment (Table 3).

**Table 3.** Multivariate Analysis – For every univariate significance of p<0.001, we conducted a multivariate analysis to reassess the significance. *Sourced from O10 (“preexisting hypertension complicating pregnancy”), which is more complete in pregnant inpatients than general chronic hypertension codes.

|  | **OR** | **95% CI** | **p-value** |
| --- | --- | --- | --- |
| **Pregnancy-Associated Conditions** |  |  |  |
| Eclampsia | 0.93 | 0.55 – 1.56 | 0.771 |
| Preeclampsia | 2.38 | 2.04 – 2.78 | <0.001 |
| Spontaneous abortion | 0.76 | 0.35 – 1.64 | 0.482 |
| Gestational edema/proteinuria (no hypertension) | 1.01 | 0.59 – 1.73 | 0.974 |
| Genitourinary tract infection in pregnancy | 0.45 | 0.31 – 0.64 | <0.001 |
| Gestational diabetes | 0.77 | 0.62 – 0.96 | 0.019 |
| Gestational hypertension (no proteinuria) | 1.66 | 1.31 – 2.11 | <0.001 |
| Malnutrition in pregnancy, childbirth and the puerperium | 1.16 | 0.63 – 2.14 | 0.637 |
| **Chronic Medical Conditions** |  |  |  |
| Hypertension (preexisting)* | 1.84 | 1.49 – 2.28 | <0.001 |
| Coronary artery disease (CAD) | 3.29 | 2.19 – 4.95 | <0.001 |
| Chronic kidney disease (CKD) | 0.56 | 0.34 – 0.90 | 0.018 |
| Chronic obstructive pulmonary disease (COPD) | 1.82 | 0.90 – 3.66 | 0.094 |
| Hyperlipidemia | 1.48 | 0.92 – 2.39 | 0.108 |
| Lupus (SLE) | 0.64 | 0.34 – 1.21 | 0.17 |
| Type 2 diabetes | 0.96 | 0.66 – 1.41 | 0.851 |
| Obesity | 0.93 | 0.75 – 1.16 | 0.51 |
| Smoking | 1.27 | 1.03 – 1.57 | 0.028 |

#### Chronic Medical Conditions

After adjustment for age, race, comorbidities, hospital characteristics, and socioeconomic factors, women with PPCM had higher odds of preexisting hypertension (OR 1.84, 95% CI 1.49–2.28, p < 0.001), coronary artery disease (OR 3.29, 95% CI 2.19–4.95, p < 0.001), and smoking (OR 1.27, 95% CI 1.03–1.57, p = 0.028). Previously significant univariate associations with COPD, hyperlipidemia, systemic lupus erythematosus, type 2 diabetes, and obesity were no longer statistically significant after adjustment (Table 3).

#### Demographics

In the adjusted model, women aged <30 still had a significantly greater risk of PPCM as those ≥30 years had lower odds of PPCM comparatively (OR 0.81, 95% CI 0.70–0.93, p = 0.004). Additionally, women from households earning <$46,000 had significantly greater odds of developing PPCM compared to households earning $46,000–$58,999 (OR 0.81, 95% CI 0.68–0.97, p = 0.022) and ≥$79,000 odds (OR 0.60, 95% CI 0.47–0.77, p < 0.001). While not significant in the univariate model, after adjustment, hospital region became a significant factor with those in the South being more likely to have PPCM compared to those in the Northeast (OR 1.36, 95% CI 1.09–1.69, p=0.006). While Black and Native American women were disproportionately represented in the PPCM group in unadjusted models, there were no significant increased risk for PPCM compared to White women after adjustment, suggesting that observed racial disparities are largely explained by differences in underlying comorbidities and socioeconomic status rather than race itself (Table 4).

**Table 4.** Multivariate Demographics.

|  | **OR** | **95% CI** | **p-value** |
| --- | --- | --- | --- |
| **Age Group** |  |  |  |
| Age<30 | REF | REF | REF |
| Age>=30 | 0.81 | 0.7 – 0.93 | 0.004 |
| **Race** |  |  |  |
| White | REF | REF | REF |
| Black | 1.07 | 0.89 – 1.27 | 0.478 |
| Hispanic | 0.71 | 0.58 – 0.88 | 0.001 |
| Asian/Pacific Islander | 0.94 | 0.68 – 1.31 | 0.71 |
| Native American | 1.41 | 0.79 – 2.53 | 0.245 |
| Others | 0.84 | 0.58 – 1.22 | 0.368 |
| **Median Household Income** |  |  |  |
| \$1 - \$45,999 | REF | REF | REF |
| \$46,000 - \$58,999 | 0.81 | 0.68 – 0.97 | 0.022 |
| \$59,000 - \$78,999 | 0.83 | 0.69 – 1.01 | 0.065 |
| \$79,000+ | 0.6 | 0.47 – 0.77 | 0 |
| **Hospital Region** |  |  |  |
| Northeast | REF | REF | REF |
| Midwest | 1.1 | 0.86 – 1.4 | 0.435 |
| South | 1.36 | 1.09 – 1.69 | 0.006 |
| West | 1.25 | 0.97 – 1.6 | 0.083 |

#### Summary

In summary, after adjustment, women with PPCM were more likely to have preeclampsia and gestational hypertension without proteinuria compared to women without PPCM. Women with PPCM were also more likely to have preexisting hypertension, coronary artery disease, and smoke compared to women without PPCM (Figure 2). Women with PPCM were additionally more likely to be <30 years old, come from lower income households, and present in hospitals located in the South.

**Figure 2.**
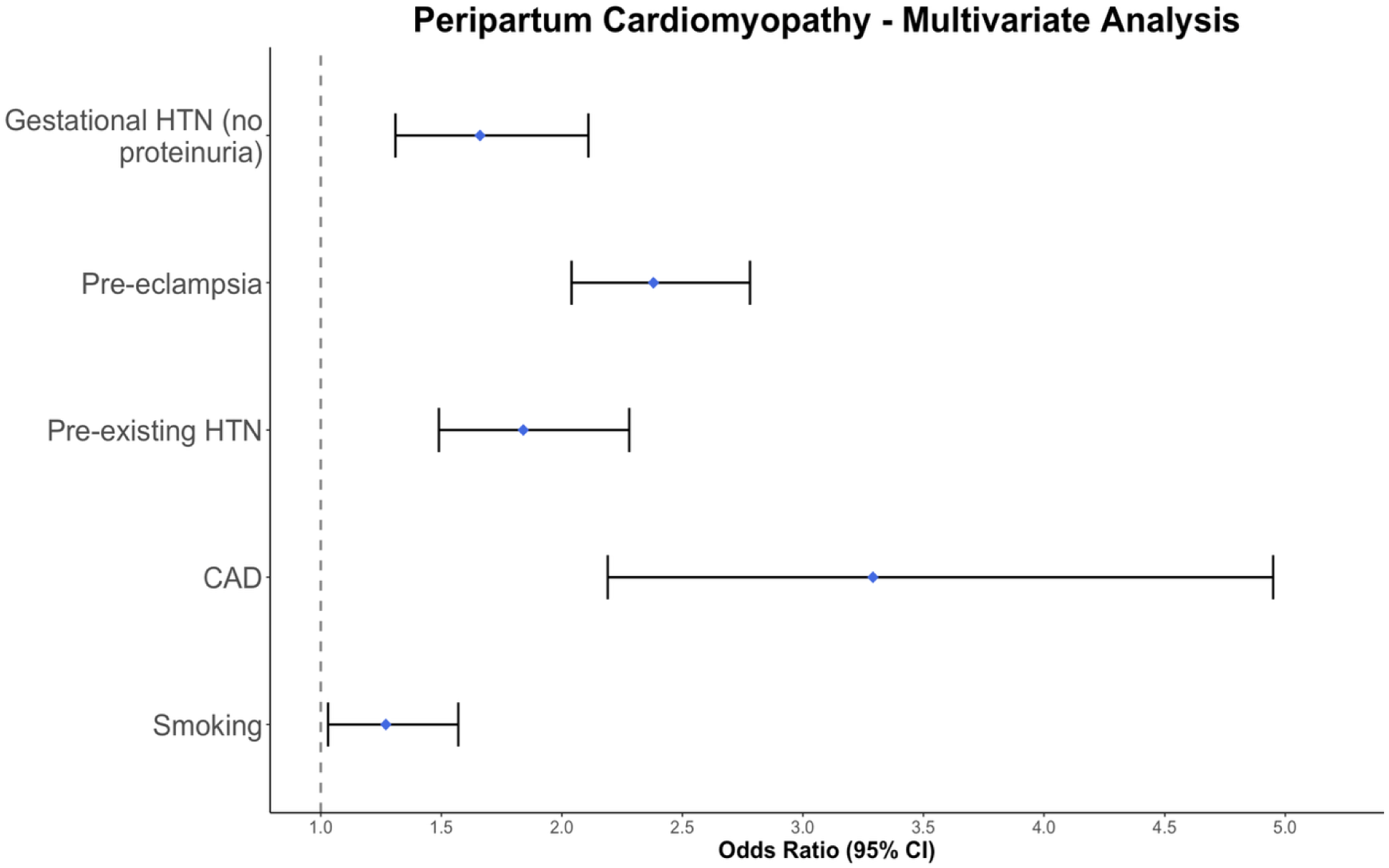
Multivariate Significant Results.

## Discussion

The univariate model identified far more associations with PPCM than the multivariate model did. The loss of significance between these models highlights the specific associations that are more likely to be impacted by more holistic variables such as socioeconomic factors and other comorbidities. The variables that lost positive significance between the models include malnutrition in pregnancy, eclampsia, spontaneous abortion, gestational edema/proteinuria without hypertension, COPD, hyperlipidemia, SLE, type 2 diabetes, obesity, genitourinary tract infection in pregnancy, gestational diabetes, and CKD. This loss of positive association is representative of variables that are confounded by other factors and has been discussed previously in the context of PPCM literature. For example, a previous study looking into the relationship between PPCM and gestational diabetes directly discusses the impact of confounding vascular risk factors ^14^. In terms of demographic data, it was found that Native American and Black racial identities were not independently associated with PPCM following adjustment for comorbid and socioeconomic variables. Importantly, this indicates that racial disparities in PPCM, as identified by the univariate models, are likely driven by inequities in underlying comorbid and socioeconomic risk factors, rather than race itself.

Several associations with PPCM persisted even after adjustment for confounders. For pregnancy-associated conditions, preeclampsia and gestational hypertension without proteinuria remained significantly associated with PPCM in the multivariate model. Among chronic medical conditions, preexisting hypertension, CAD, and smoking maintained significant positive associations. Demographically, there remained an association between PPCM and income even after adjustment as those in the highest income bracket were significantly less likely than those in the lowest income bracket to have PPCM. These persistent associations suggest that, even after accounting for other socioeconomic factors and comorbid conditions, these variables are independently related to PPCM risk. As previously mentioned, a recent meta-analysis identified obesity, multiparity, gestational hypertension, diabetes, and preeclampsia as risk factors for PPCM with a history of smoking not being associated with increased risk for PPCM ^12^. Our study provides further support for preeclampsia and gestational hypertension being risk factors for PPCM, but it calls into question whether obesity and diabetes are truly independently associated with PPCM risk and highlights the need for more research surrounding smoking as a risk factor. It is also important to note that the meta-analysis pulls in papers globally, whereas our data is pulled solely from the U.S. This also points to the need for additional research surrounding global differences in PPCM risk.

In terms of possible regional differences within the U.S., we found that after adjustment, hospital region became a significant factor, with those in the South being more likely to have PPCM compared to those in the Northeast (OR 1.36, 95% CI 1.09–1.69, p=0.006). On a global scale, there are studies which show that the incidence of PPCM is highest in countries with tropical climates and distinct wet and dry seasons. It is postulated that this may be a result of autoimmune mechanisms that evolved to protect pregnant women in tropical regions ^5^. Whether climate or latitude meaningfully influences PPCM risk within the United States remains unclear but deserves further investigation into whether environmental factors, including temperature, humidity, or infectious exposures, may still play a role in modulating inflammatory pathways that may predispose certain individuals to PPCM. Furthermore, from a regional socioeconomic standpoint, our data corroborates with a study that compared the impacts of Social Vulnerability Indices (SVI) on mortality from PPCM amongst US census regions with the highest being in the South and the lowest being in the Northeast ^15^. More research must be done to determine the degree and manner in which environmental and socioeconomic factors may affect rates of PPCM and its geographical distribution.

One unique finding that was significant in both univariate and multivariate models was related to the age of patients with and without PPCM. Although the mean age of patients with PPCM was slightly higher than those without PPCM (30.40 vs. 29.09 years), there was a disproportionate number of patients <30 years old with PPCM (49.72%) compared to the overall number of patients <30 years old (41.37%). At first glance, this may seem contradictory, but this likely reflects differences in the denominator populations. Older pregnant women have more frequent hospitalizations (58.63%) likely for a wider variety of complications, making PPCM a smaller fraction of their total admissions, whereas younger women have fewer admissions overall (41.37%), likely making PPCM a proportionally larger share. Additional contributors may include more frequent inpatient monitoring of “high-risk” pregnancies in older women, further diluting the relative PPCM proportion, and possible diagnostic vigilance in younger women when symptoms are atypical for their age and thus more readily identified as not being physiological changes related to pregnancy.

Notably, CAD had the strongest association with PPCM in both the univariate (OR 125.86) and multivariate (OR 3.29) models, suggesting it may be the single most impactful comorbidity related to PPCM. Interestingly, prior work has shown that CAD in the context of PPCM is linked with adverse events such as STEMI, yet paradoxically protective against preeclampsia ^16^. It is important to note that largest association across all data points was in maternal mortality (OR 164.41). It is quite striking to consider that women with PPCM are more than 164 times more likely to die in the hospital compared to women without PPCM. For both CAD and maternal mortality, the magnitude of the odds ratio is partly inflated by the rarity of these events in women without PPCM, but the associations remain clinically compelling and underscore the urgent need for further research to improve the treatment of this condition.

Overall, these findings underscore the complexity of PPCM’s etiology, where a mosaic of socioeconomic determinants, pregnancy-related complications, and underlying comorbid conditions collectively influence disease risk. However, certain associations, such as CAD, preeclampsia, gestational hypertension without proteinuria, preexisting hypertension, smoking, and lower-income households, appear independently associated with increased risk of PPCM. We hope that through providing these associations, healthcare providers will be able to better assess the risk of PPCM and distinguish it from physiological symptoms of pregnancy earlier on in its course to reduce the overall mortality associated with it.

## Limitations

Our study evaluated only patients who were admitted to a hospital and therefore cannot represent all pregnancies that will lead to an overestimation of the PPCM incidence. We used ICD-10 codes with inherent limitations. We cannot rule out other rare conditions that can also contribute to the higher prevalence of PPCM in our study. This is a retrospective study requiring confirmations in prospective studies.

## Conclusion

PPCM is associated with higher mortality. It is independently associated with preexisting cardiovascular risk factors. However, apparent racial disparities and association with socioeconomic factors disappear after adjustment, suggesting that those associations primarily reflect comorbidities and social determinants and are not independent risk.

## Ethical Statement

The NIS database is publicly available, without any patient identifiers, exempt from IRB or obtaining informed consent.

## Data Sharing

The NIS database is publicly available upon purchase

## Data Availability

The NIS database is publicly available uppon purchase

---

**Table S1.** ICD-10 Codes.

| Condition | ICD-10 Codes |
| --- | --- |
| Peripartum cardiomyopathy | O90.3 |
| Weeks of gestation | Z3A |
| Preexisting hypertension complicating pregnancy, childbirth and the puerperium | O10 |
| Gestational [pregnancy-induced] edema and proteinuria without hypertension | O12 |
| Gestational [pregnancy-induced] hypertension without significant proteinuria | O13 |
| Preeclampsia | O14, O11 |
| Eclampsia | O15 |
| Hemorrhage in early pregnancy | O20 |
| Excessive vomiting in pregnancy | O21 |
| Infections of genitourinary tract in pregnancy | O23 |
| Diabetes mellitus in pregnancy, childbirth, and the puerperium | O24 |
| Malnutrition in pregnancy, childbirth and the puerperium | O25 |
| Spontaneous abortion | O03 |
| Rheumatoid arthritis | M05, M06 |
| Scleroderma (systemic sclerosis) | M34 |
| Systemic connective tissue disorders | M35 |
| Gout | M10, M1A |
| Psoriatic arthritis | L40.50 |
| Cryoglobulinemia | D89.1 |
| Polyarteritis nodosa and related conditions | M30 |
| Reynaud's syndrome | I73.0 |
| Buerger's disease (thromboangiitis obliterans) | I73.1 |
| Ankylosing spondylitis | M45 |
| Antiphospholipid antibody syndrome | D68.61, D68.312 |
| Third degree AV block | I44.2 |
| Endocarditis (Mitral, Tricuspid, Multiple valves, Infective acute/subacute, Acute/subacute, Aortic valve, Pulmonary valve) | I05.9, I07.9, I08.9, I33.0, I33.9, I35.8, I37.8 |
| Coronary Artery Disease | I2101,I2102,I2109,I2111,I2119,I2121,I2129,I213,I219,I21A1,I21A9,I220,I221,I225,I229,I214,I222,I251,I2510,I2511,I25110,I25111,I25112,I25118,I25119,I255,I256,I2582,I2583,I2584,I2589,I259 |
| Lupus | M32.10,M32.11,M32.12,M32.14,M32.15,M32.19,M32.8,M32.9 |

